# Clinical and population genomic epidemiology of invasive group A streptococcus in Scotland, 2014-2024

**DOI:** 10.64898/2026.07.13.26357965

**Authors:** Stephen B. Beres, Davide Pagnossin, Randall J. Olsen, S. Wesley Long, Edward A. Graviss, Thomas Williams, Ross Langley, Andrew Smith, James M. Musser

## Abstract

**Objectives:** Following the COVID-19 pandemic, multiple countries reported a surge in invasive group A streptococcus (iGAS) infections. Posited explanations include reduced population immunity, increased respiratory virus co-infection, and emergence of hypervirulent GAS clones. To assess the relative contribution of these factors, we analyzed the epidemiology and genomics of 3,408 iGAS infections in Scotland.

**Methods:** National surveillance data from 2014–2024 were analyzed to characterize iGAS incidence. Hybrid whole genome sequencing was used to comprehensively genetically characterize 404 *emm*1 isolates collected from invasive and tonsillitis infections.

**Results:** iGAS incidence markedly increased in late 2022 and early 2023, disproportionately affecting children and older adults. This surge was not associated with a proportional increase in bacteremia but did coincide with increased influenza and respiratory syncytial virus infections. Genomic analyses found that *emm*1 post-pandemic isolates were not genetically distinct from pre-pandemic isolates in genome-wide polymorphisms, accessory genes including virulence and antimicrobial resistance determinants, mobile genetic elements, or chromosomal structural variants.

**Conclusions:** The post-pandemic iGAS surge in Scotland was not associated with emergence of a novel hypervirulent *emm*1 clone. Instead, the epidemiologic and population genomic findings are consistent with increased host susceptibility following reduced pathogen exposure during the pandemic and increased respiratory virus co-infection as predominant contributing factors.

**Highlights:**

- Post-COVID-19 iGAS surged disproportionately affecting children and the elderly
- The iGAS surge was associated with an overall decreased odd of bacteremia
- Peaks in influenza and RSV activity coincided with the post-pandemic iGAS surge
- Emm1 was the most prevalent *emm* type, accounting for 56% of the iGAS surge isolates
- Emergence of a more virulent *emm*1 clone was not supported by genomic analyses

## Introduction

Recent studies from multiple countries have described the epidemiology of invasive GAS infections reporting a marked increase incidence in late 2022 and early 2023 (Abo et al. 2023; Beres et al. 2024; Cobo-Vazquez et al. 2023; de Gier et al. 2023; Golden et al. 2024; Gouveia et al. 2023; Guy et al. 2023; Johannesen et al. 2023; Lassoued et al. 2023; Nygaard et al. 2024; Valcarcel Salamanca et al. 2024). This increase followed a period of historically low activity during most of 2020 and 2021 when many countries had implemented non-pharmaceutical interventions (NPIs) to stem transmission of the COVID-19 virus.

Epidemiological patterns similar to those of iGAS have also been reported for several other respiratory pathogens, including *Streptococcus pneumoniae*, *Haemophilus influenzae*, *Mycoplasma pneumoniae*, respiratory syncytial virus (RSV), and influenza virus, with reduced incidence during the COVID-19 NPIs followed by a rapid resurgence after their relaxation (Burrell, Saravanos, and Britton 2025; Meyer Sauteur et al. 2024; Singer et al. 2024). Although these epidemiological changes have been documented in countries across Europe, North America, and Oceania, data from many regions of Africa, Asia, China, and South America remain scarce, limiting the generalizability of these observations at a global level.

The post-pandemic increase in iGAS infections, although observed across all ages, was frequently reported to disproportionately affect children and was associated with unusually high numbers of respiratory manifestations, including pneumonia and empyema (de Gier et al. 2023; Guy et al. 2023; Nygaard et al. 2024). Temporal and clinical associations of iGAS with respiratory viral infections have repeatedly been described, suggesting that predisposing viral infections may have contributed to increased iGAS incidence (de Gier et al. 2024; Engstrom et al. 2023; Guy et al. 2023; Lassoued et al. 2023; Lees et al. 2024). Most studies reported a predominance of *emm*1 isolates during the post-pandemic iGAS surge (Gouveia et al. 2023; Rodriguez-Ruiz et al. 2023; Vieira et al. 2024), however, it is important to note that, historically, *emm*1 was already the most common *emm* type associated with iGAS disease in high-income countries independent of the pandemic (Steer et al. 2009). Moreover, while some studies identified expansion of the more recently emerged M1 United Kingdom (M1_UK_) lineage in association with the iGAS infection surge (Beres et al. 2024; Davies et al. 2023; Gouveia et al. 2023), others reported continued predominance of the predecessor M1 global (M1_GLB_) lineage (Cipolla et al. 2025; Ramirez de Arellano et al. 2024) or other *emm* types across different settings (Davies et al. 2025; Golden et al. 2024; van der Putten et al. 2023), demonstrating that the geographically widespread increased iGAS incidence observed post-pandemic was polyclonal in nature.

Although factors contributing to the post-pandemic surge in iGAS disease are not completely understood, a review across investigations has posited three main hypotheses (Flamant et al. 2025). First, reduced herd immunity -- reduced exposure to respiratory pathogens (both viral and bacterial) during the period of COVID-19 NPIs may have resulted in reduced population immunity, often referred to as "immunity debt", increasing susceptibility to GAS and other common respiratorially transmitted pathogens. Second, predisposing viral infections -- increased incidence of preceding or coincident respiratory viral infections may have contributed to increased GAS infection incidence and severity. Third, virulent clone emergence -- the emergence and expansion of GAS clones with increased fitness, transmissibility, and/or virulence may have contributed to increased GAS infection. Among these three hypotheses, the emergence of more virulent clones is the most speculative as it lacks supporting comparative genomic data (Flamant et al. 2025).

Herein, we present 11 years (2014–2024) of national reference laboratory surveillance data from Scotland to comprehensively characterize the epidemiology of iGAS infections in correlation with the COVID-19 pandemic. Additionally, we evaluate how well the Scotland data comport with the three main hypotheses proposed to explain the unprecedented post-pandemic iGAS surge. First, to assess the potential impact of reduced population immunity, incidence rates (IRs) were evaluated relative to specific age groups, given that the young are immunologically naive and the elderly experience age-related immunosenescence. Second, to evaluate the role of predisposing viral infections, we assessed iGAS incidence in relation to the population-level incidence of circulating respiratory viruses. Third, to directly evaluate the virulent clone hypothesis, we applied hybrid whole-genome sequencing (WGS), combining short- and long-read technologies, to comprehensively genetically characterize a large collection of *emm*1 isolates (*n* = 404) obtained from both invasive and non-invasive infections. This generated a high-resolution population genomic dataset that enabled robust phylogenetic and genome-wide association (GWAS) analyses to determine if the emergence and expansion of a genetically distinct, more transmissible and/or virulent *emm*1 clone temporally correlated with and contributed to the post-pandemic iGAS surge.

## Materials and methods

### Public Health Scotland surveillance of GAS infections 2014-2024

Invasive GAS infections are notifiable in Scotland under the Public Health (Scotland) Act 2008. Culture and PCR-positive iGAS specimens from all Scottish laboratories are reported to Public Health Scotland (PHS) using the Electronic Communication of Surveillance in Scotland (ECOSS) system. From the diagnostic and reference laboratory perspective, isolates are referred based on UK guidance (UKHSA 2023), which includes specimens from normally sterile body sites or non-sterile sites if in combination with a severe clinical presentation: streptococcal toxic shock syndrome, necrotizing fasciitis, pneumonia, septic arthritis, meningitis, peritonitis, osteomyelitis, myositis, and puerperal sepsis.

### Scottish GAS reference laboratory surveillance of GAS infections

Clinical iGAS samples are submitted to Scottish Microbiology Reference Laboratory (SMiRL) from routine microbiology diagnostic laboratories for further analysis and typing. All isolates are sub-cultured onto Columbia horse blood agar plates (Oxoid) and incubated at 37°C for 24 h in an atmosphere of 5 % CO_2_. The isolates are phenotypically identified by colony morphology, beta-hemolysis and Lancefield group identification using a Prolex Strep Grouping Latex Test Kit. DNA is prepared and e*mm* typing is performed as described (CDC 2024), using an AB 3500xl sequencer. The sequences are manually edited using CLC Main Workbench software, and the FASTA files are uploaded to the BLAST 2.0 server on the CDC website (CDC 2024). iGAS isolate data for this study were extracted from the Laboratory Information Management System into excel spreadsheets, and duplicate isolates were removed.

### Viral respiratory tract infections

National data on influenza and respiratory syncytial virus (RSV) hospital admissions over the 11-year study period were provided by PHS from the SMR01 database, which receives notifications through ECOSS. Hospital admissions were defined as a positive influenza or RSV test within 14 days before, or two days after, hospital admission.

### Statistical analyses

All incidence rates are annualized, presented as incidence per 100,000 population per year. Specifically, quarterly incidences were multiplied by four to estimate the annual incidence. Population sizes were based on mid-year population estimates obtained from the National Records of Scotland (nrscotland.gov.uk).

Changes in infection incidence over time were tested using simple linear regression trend analysis using Prism v11. The significance of change in IRs relative to the pandemic intervals was tested by Poisson regression in R v4.5.2.

To test whether the post-pandemic surge affected age groups proportionally, a generalized linear model (GLM) with a Poisson distribution and log-link function was fitted. The model included age group, time period in relation to the COVID-19 pandemic, and their interaction to evaluate whether changes in incidence varied by age. The log of person-years at risk was included as an offset to model incidence rates rather than case counts. This modelling approach was applied to both iGAS cases overall and blood culture–positive isolates.

To determine if the total number of iGAS infections during the pandemic and post-pandemic periods differed from that expected based on pre-pandemic norms, an expected-versus-observed analysis was conducted. The mean incidence rate derived from the 25-quarter pre-pandemic period was used to predict the expected number of cases for the subsequent 19 quarters, encompassing both the pandemic and post-pandemic periods. Expected case counts were adjusted for changes in population size using mid-year population estimates, thereby accounting for differences in person-time at risk. Observed and expected case counts were compared using chi-squared (χ²) goodness-of-fit tests. This analysis was performed for both all iGAS cases and blood culture–positive iGAS cases, including stratified analyses by age group.

Linear regression was also used to estimate the mean proportion of blood culture– positive isolates among all invasive isolates during the pre-pandemic period. Logistic regression was then used to compare odds of blood culture positivity during the post-pandemic surge with pre-pandemic levels. The same approach was applied to the proportion of *emm*1 blood culture–positive isolates among all invasive *emm*1 cases.

### Selection of emm1 isolates for WGS

Historical SMiRL records were used to retrieve iGAS isolates of all *emm* types collected between 2014 and 2024. WGS genetic characterization was done for a subset of 404 *emm*1 isolates. This *emm* type focus was prompted by *emm*1 being the most prevalent cause of iGAS infections throughout the surveillance study period, and in particular being predominant during the late 2022 - early 2023 surge in iGAS infections. The selected *emm*1 WGS isolates included 25.8% (232 /898) of all *emm*1 invasive infection isolates collected from 2014-2024, and virtually all (172/179) *emm*1 tonsillitis isolates collected via enhanced surveillance conducted in late 2022 prompted by the pediatric surge in iGAS infections. Selected *emm*1 isolates included all age groups and pandemic intervals but disproportionately favored pediatric isolates and isolates of infections that occurred during the surge in iGAS infections. Isolates were subcultured to ensure purity and sent to the Houston Methodist Research Institute for WGS.

### Population genomic analyses

Whole genome sequencing genetic characterization of *emm*1 isolates was performed using a combination of Illumina paired-end short-read sequencing and Oxford Nanopore Technologies long-read sequencing multiplexed libraries (Supplementary Table S1) as previously described (Beres et al. 2023). Hybrid genome assemblies were generated using Hybracter v0.13.0, resulting in complete closed genome sequences for 399 of 404 isolates. For the remaining five isolates, incomplete partial genome assemblies were generated from Illumina short-reads using SPAdes v4.2.0. Genome assemblies were annotated with MGAS2221 as reference (accession: ASM1257226v1) using Prokka v1.15.6.

Single nucleotide polymorphisms (SNPs) were identified relative to the MGAS2221 genome using the MUMmer v4.1.0 dnadiff function. Phylogenies were inferred from core genome SNPs by Maximum-Likelihood using IQ-TREE v2.4.0 under a general time-reversible (GTR) model of nucleotide substitution. Lineage-defining SNPs were used to classify isolates into M1_GLB_ (Global), M1_INT_ (Intermediate) and M1_UK_ (United Kingdom) lineages (Lynskey et al. 2019). Emm type and multilocus sequence types were determined using Emmtyper v0.2.0 and FastMLST v0.0.19. Recombination within the M1 population was assessed using Gubbins v3.4.3.

Genome-wide association studies (GWAS) were conducted using PYSEER v1.4.0 on M1_UK_ isolates (*n* = 380) pangenome, comparing pre-pandemic and post-pandemic isolates. Significantly non-randomly distributed small genetic variation (ex. SNPs and indels) for the pangenome was characterized genome-wide using unitigs determined using unitig-caller, and for the accessory portion of the pangenome using variably present genes determined using Panaroo v1.5.1.

Antimicrobial resistance genes were identified using AMRFinderPlus 4.0.19. The presence of mobile genetic elements (MGEs) was inferred through detection of integrase genes and phage-associated virulence factors using a previously published reference database (Kachroo et al. 2019).

GAS chromosomes can differ in architecture due to large structural variants (SVs), chromosomal inversion and rearrangement events. Large SVs were evaluated among the 399 complete closed *emm*1 genome assemblies by aligning them pairwise to the core genomes of M1_GLB_ reference strains MGAS5005 and MGAS2221 using minimap2 v2.28.

### Ethics

iGAS isolates were investigated in accordance with the Public Health (Scotland) Act 2008. Public Health Scotland is mandated to process data relating to notifiable diseases, health risk states, notifiable organisms, and public health investigations. As such, individual patient consent was not required. Information governance approval for the sharing of patient demographic data for the purpose of this investigation was granted by the Scottish Information Governance, Public Benefit and Privacy Panel for Health and Social Care (application reference 2223-0204 Smith). For GAS tonsillitis isolates, information governance approval for sharing of patient demographic data was granted by the local Caldicott Guardian for NHS Greater Glasgow and Clyde. No patient-identifiable information is included in this paper.

## Results

### Impact of the COVID-19 pandemic on iGAS infection incidence

A total of 3,408 iGAS cases were recorded in Scotland for the 11-year surveillance study period (2014 to 2024). Over the 25 quarters preceding the COVID-19 pandemic (2014-Q1 to 2020-Q1, 6.25 years), the all-ages iGAS IRs were stable (Fig. 1A and Table 1), with no significant trend up or down (linear regression: slope = 0.056, p = 0.726), establishing a pre-pandemic norm. During the 8-quarter COVID-19 pandemic interval (2020-Q2 to 2022-Q1, 2 years), during which the NPIs (i.e. masking, social distancing, closure of non-essential businesses, prohibition of large public gatherings, etc.) were implemented, the median iGAS IR significantly decreased by 71% relative to pre-pandemic (IRR = 0.29, p < 0.001). Following the lifting of NPIs, iGAS IRs dramatically surged over late 2022 and early 2023 (2022-Q4 to 2023-Q2, 0.75 years), reaching a median nearly three times higher than pre-pandemic (IRR = 2.77, p < 0.001). Over the next 6 quarters following the surge (2023-Q3 to 2024-Q4, 1.5 years), the iGAS IRs dropped back to levels no longer significantly different from pre-pandemic (IRR = 1.10, p = 0.052). In contrast to the dramatic fluctuations in iGAS IRs over the study period, the Scottish population remained stable, not varying in correlation with the COVID-19 pandemic intervals (Supplementary Figure S1).

**Fig. 1.**
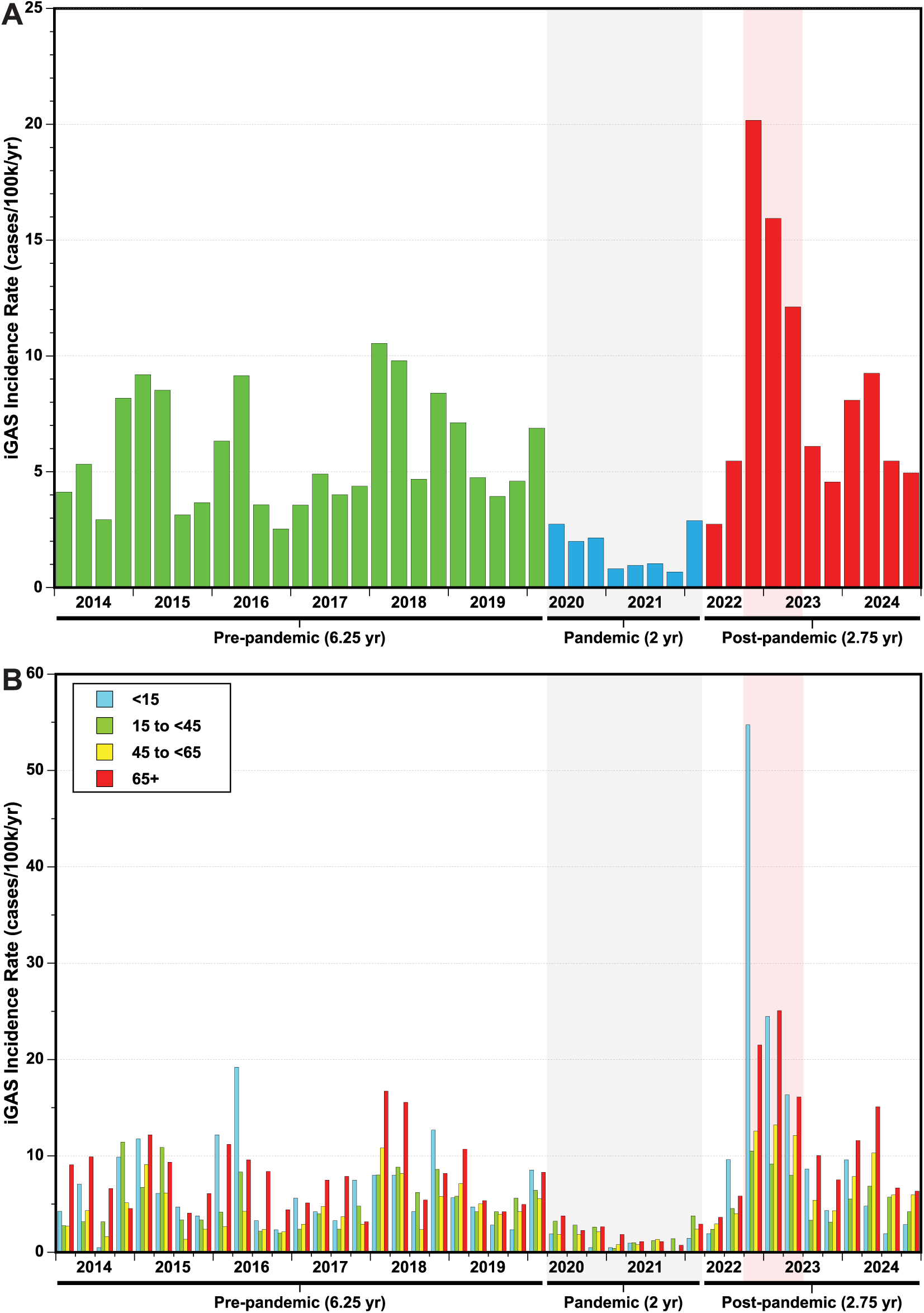
(**A**) Invasive group A streptococcus quarterly incidence rates of in Scotland, 2014–2024. (**B**) Invasive group A streptococcus quarterly incidence rates stratified by age group. COVID-19 pandemic and post-pandemic surge intervals are highlighted in gray and red respectively.

**Table 1.** Incidence rates, linear trend, and Poisson regression analyses of invasive group A streptococcus infections in Scotland by age group and COVID-19 pandemic intervals.

| Age group | COVID-19 Interval | Median IR [IQR] | IRR (95% CI) | p-value |
| --- | --- | --- | --- | --- |
| All ages | Pre-pandemic | 4.7 [3.9-8.2] | linear regression: m = 0.056, p = 0.726 |  |
|  | Pandemic | 1.5 [0.9-2.3] | 0.29 (0.25-0.33) | < 0.001 |
|  | Post-pandemic | 6.0 [5.2-10.6] | 1.48 (1.38-1.59) | < 0.001 |
|  | Surge | 15.8 [13.9-17.9] | 2.77 (2.53-3.02) | < 0.001 |
|  | Post-surge | 5.7 [5.0-7.5] | 1.10 (1.00-1.21) | 0.052 |
| <15 years | Pre-pandemic | 5.6 [3.8-8.0] | linear regression: m = 0.035, p = 0.707 |  |
|  | Pandemic | 0.5 [0.0-1.1] | 0.10 (0.05-0.17) | < 0.001 |
|  | Post-pandemic | 8.6 [3.6-13.0] | 1.95 (1.66-2.27) | < 0.001 |
|  | Surge | 24.5 [20.4-39.6] | 4.90 (4.11-5.82) | < 0.001 |
|  | Post-surge | 4.6 [3.2-7.7] | 0.82 (0.63-1.06) | 0.145 |
| 15 to <45 years | Pre-pandemic | 4.2 [3.2-6.7] | linear regression: m = 0.063, p = 0.657 |  |
|  | Pandemic | 2.0 [1.2-2.9] | 0.39 (0.31-0.48) | < 0.001 |
|  | Post-pandemic | 5.5 [3.8-7.4] | 1.08 (0.95-1.21) | 0.248 |
|  | Surge | 9.1 [8.6-9.8] | 1.73 (1.44-2.07) | < 0.001 |
|  | Post-surge | 4.9 [3.5-5.7] | 0.90 (0.75-1.07) | 0.255 |
| 45 to <65 years | Pre-pandemic | 4.2 [2.7-5.6] | linear regression: m = 0.192, p = 0.227 |  |
|  | Pandemic | 1.6 [0.8-1.9] | 0.31 (0.22-0.42) | < 0.001 |
|  | Post-pandemic | 6.0 [4.9-11.2] | 1.72 (1.49-1.99) | < 0.001 |
|  | Surge | 12.6 [12.3-12.9] | 2.83 (2.33-3.42) | < 0.001 |
|  | Post-surge | 6.0 [5.5-7.4] | 1.48 (1.23-1.79) | < 0.001 |
| ≥65 years | Pre-pandemic | 7.9 [5.1-9.6] | linear regression: m = 0.034, p = 0.757 |  |
|  | Pandemic | 2.1 [1.1-2.7] | 0.26 (0.19-0.35) | < 0.001 |
|  | Post-pandemic | 10.0 [6.5-15.6] | 1.48 (1.29-1.70) | < 0.001 |
|  | Surge | 21.5 [18.8-23.3] | 2.63 (2.21-3.12) | < 0.001 |
|  | Post-surge | 8.8 [6.9-11.2] | 1.20 (1.00-1.43) | 0.041 |

The annualized incidence rate (IR) per 100,000 population is presented as the median and interquartile range [IQR] of the quarterly rates within each defined interval. To establish the pre-pandemic norm, simple linear regression trend analysis was performed on the quarterly IRs over the 6.25-year pre-pandemic interval, yielding the slope (m) and statistical significance (p-values) to confirm there was no significant upward or downward trend prior to the pandemic. Incidence rate ratios (IRR) with 95% confidence intervals (CI) and statistical significance (p-values) were calculated using a generalized linear model for Poisson regression. To adjust for varying population sizes and exposure intervals, the model utilized the number of iGAS cases as the dependent variable, the COVID-19 pandemic interval as the independent categorical variable, and the natural logarithm of the population-time at risk (person-years) as the offset. The IRRs represent the relative risk of iGAS infection during the pandemic, post-pandemic, surge, and post-surge intervals compared to the historic baseline norm established during the pre-pandemic reference interval.

The demographic distribution of iGAS infections was evaluated across four age groups, namely: <15, 15 to <45, 45 to <65, and ≥65 years (Fig. 1B). Prior to the COVID-19 pandemic, the quarterly iGAS IRs across all age groups were stable, showing no trends up or down (all p-values are > 0.05), establishing age group specific pre-pandemic norms (Table 1). During the pandemic interval, the iGAS IR significantly decreased across all age groups (all IRR ≤ 0.39, p < 0.001). Following the lifting of pandemic restrictions, the iGAS IR increased significantly across all age groups over the surge interval (all IRR ≥ 1.73, p < 0.001). This increase was greatest for the young, with the <15 group experiencing a nearly 5-fold increase (IRR = 4.90, p < 0.001). The ≥65 group experienced the second largest increase, approximately half that of the <15 (IRR = 2.63, p < 0.001). In the post-surge interval, iGAS IRs for the <15 and 15 to <45 groups dropped back to levels no longer significantly different from the pre-pandemic baselines (p = 0.145 and p = 0.255, respectively). Conversely, post-surge iGAS IRs for the 45 to <65 and ≥65 groups remained elevated compared to the pre-pandemic baselines, although the magnitude of the post-surge increase was less than 50% (IRR = 1.48, p < 0.001; and IRR = 1.20, p = 0.041, respectively).

The iGAS disease burden was next evaluated to determine whether the increased risk during the surge interval was distributed equivalently across the age groups, or if any age groups were disproportionately affected. The median IR for individual age groups were compared against the median IR across all age groups (Supplementary Table S2). During the surge, the iGAS disease burden disproportionately affected the <15 group, with the young experiencing a relative iGAS risk nearly twice that of the overall Scottish median (IRR = 1.97, p < 0.001). While the ≥65 group also experienced a higher iGAS risk during the surge interval, the increase was a relatively modest 29% (IRR = 1.29, p = 0.002). Conversely, the 15 to <45 and 45 to <65 groups experienced lower iGAS risks compared to the overall median (IRR = 0.60, p < 0.001; and IRR = 0.79, p = 0.006, respectively).

### Impact of the COVID-19 pandemic on iGAS infection severity

To evaluate if the pandemic caused a shift towards more severe iGAS infections, blood culture-positive (BC+) iGAS cases were utilized as a surrogate marker for more severe disseminated systemic infections. Of the 3,408 iGAS cases, 1,791 (53%) were BC+. Considering all iGAS cases independent of age (i.e. all-ages), pre-pandemic the BC+ iGAS quarterly IRs were stable, with no trends up or down (p > 0.05) (Fig. 2, Table 2). During the pandemic interval, the all-ages overall median BC+ IR decreased 3.2-fold relative to pre-pandemic (IRR = 0.31, p < 0.001). During the surge interval, the median BC+ IR increased 2.4-fold relative to pre-pandemic (IRR = 2.44, p < 0.001). Although the BC+ iGAS IR transiently increased following the lifting of the pandemic NPIs, the proportion of iGAS cases BC+ did not (Supplementary Figure S2). To the contrary, the mean proportion of BC+ iGAS cases during the surge interval (49% ± 14%) was lower than pre-pandemic (55% ± 10%) (p = 0.005), although the medians were nearly identical. Consistent with this lower %BC+ iGAS cases, the odds of an iGAS infection presenting as BC+ decreased during the surge interval relative to the pre-pandemic norm (OR = 0.77, p = 0.005).

**Fig. 2.**
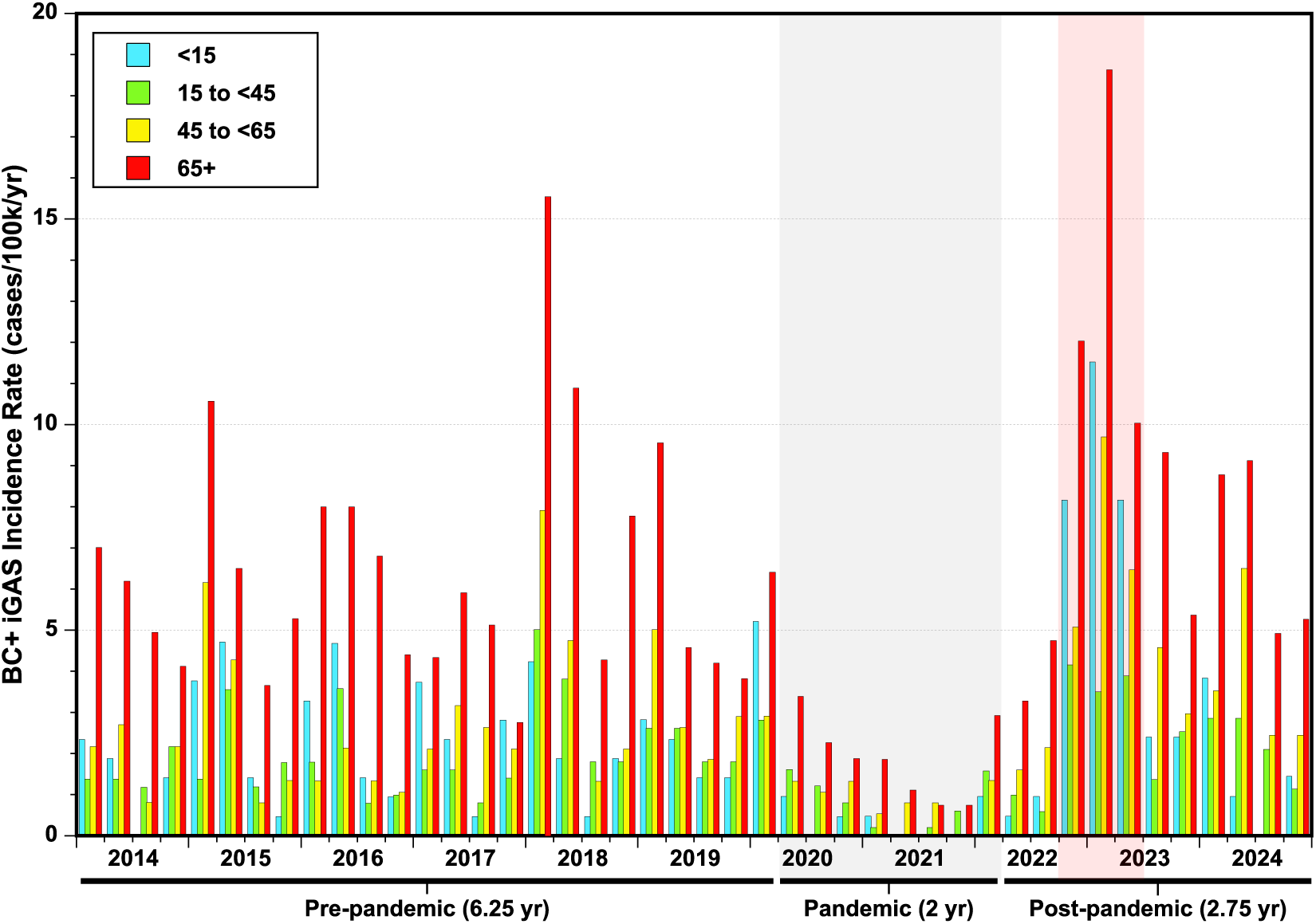
Blood culture-positive invasive group A streptococcus quarterly incidence rates in Scotland, 2014–2024, stratified by age group. Pandemic and post-pandemic surge intervals are highlighted in gray and red, respectively.

**Table 2.** Incidence rates, linear trend, and Poisson regression analyses of blood culture-positive invasive group A streptococcus cases in Scotland by age group and COVID-19 pandemic intervals.

| Age group | COVID-19 Interval | Median IR [IQR] | IRR (95% CI) | p-value |
| --- | --- | --- | --- | --- |
| All ages | Pre-pandemic | 2.7 [2.2-3.9] | linear regression: m = 0.032, p = 0.424 |  |
|  | Pandemic | 0.9 [0.4-1.3] | 0.31 (0.25-0.38) | < 0.001 |
|  | Post-pandemic | 4.0 [2.4-5.7] | 1.39 (1.26-1.53) | < 0.001 |
|  | Surge | 6.6 [6.5-8.0] | 2.44 (2.14-2.76) | < 0.001 |
|  | Post-surge | 3.6 [2.6-4.3] | 1.15 (1.01-1.30) | 0.037 |
| <15 years | Pre-pandemic | 1.9 [1.4-3.3] | linear regression: m = -0.018, p = 0.666 |  |
|  | Pandemic | 0.2 [0.0-0.6] | 0.16 (0.06-0.32) | < 0.001 |
|  | Post-pandemic | 2.4 [1.0-6.0] | 1.60 (1.21-2.11) | < 0.001 |
|  | Surge | 8.2 [8.2-9.8] | 4.05 (2.94-5.51) | < 0.001 |
|  | Post-surge | 1.9 [1.1-2.4] | 0.80 (0.50-1.23) | 0.331 |
| 15 to <45 years | Pre-pandemic | 1.8 [1.4-3.3] | linear regression: m = 0.039, p = 0.186 |  |
|  | Pandemic | 0.7 [0.2-1.3] | 0.38 (0.26-0.55) | < 0.001 |
|  | Post-pandemic | 2.5 [1.3-3.2] | 1.17 (0.95-1.44) | 0.144 |
|  | Surge | 3.9 [3.7-4.0] | 1.90 (1.42-2.51) | < 0.001 |
|  | Post-surge | 2.3 [1.5-2.8] | 1.06 (0.80-1.38) | 0.672 |
| 45 to <65 years | Pre-pandemic | 2.3 [1.3-2.9] | linear regression: m = 0.040, p = 0.409 |  |
|  | Pandemic | 0.9 [0.7-1.3] | 0.33 (0.22-0.48) | < 0.001 |
|  | Post-pandemic | 3.5 [2.4-5.8] | 1.59 (1.31-1.93) | < 0.001 |
|  | Surge | 6.5 [5.8-8.1] | 2.61 (2.02-3.34) | < 0.001 |
|  | Post-surge | 3.2 [2.6-4.3] | 1.38 (1.07-1.76) | 0.011 |
| ≥65 years | Pre-pandemic | 5.9 [4.3-7.8] | linear regression: m = -0.007, p = 0.936 |  |
|  | Pandemic | 1.9 [1.0-2.4] | 0.29 (0.21-0.39) | < 0.001 |
|  | Post-pandemic | 8.8 [5.1-9.7] | 1.29 (1.10-1.51) | 0.001 |
|  | Surge | 12.0 [11.0-15.3] | 2.11 (1.71-2.59) | < 0.001 |
|  | Post-surge | 7.1 [5.3-9.0] | 1.11 (0.90-1.35) | 0.321 |

The annualized incidence rate (IR) per 100,000 population for severe (BC+) infections is presented as the median and interquartile range [IQR] of the quarterly rates within each defined interval. To establish the pre-pandemic norm, simple linear regression trend analysis was performed on the quarterly IRs over the 6.25-year pre-pandemic interval, yielding the slope (m) and statistical significance (p-values) to confirm there was no significant upward or downward trend prior to the pandemic. Incidence rate ratios (IRR) with 95% confidence intervals (CI) and statistical significance (p-values) were calculated using a generalized linear model for Poisson regression. To adjust for varying population sizes and exposure intervals, the model utilized the number of BC+ iGAS cases as the dependent variable, the COVID-19 pandemic interval as the independent categorical variable, and the natural logarithm of the population-time at risk (person-years) as the offset. The IRRs represent the relative risk of a severe BC+ infection during the pandemic, post-pandemic, surge, and post-surge intervals compared to the historic baseline norm established during the pre-pandemic reference interval.

Stratified across the individual age groups, the BC+ iGAS IR significantly decreased during the pandemic interval and increased during the surge interval for all groups (Fig. 2, Table 2). BC+ iGAS IRs were stable over the pre-pandemic interval for each group. Pre-pandemic, the BC+ IR was highest for the ≥65 group. During the surge, the BC+ IR remained highest in the ≥65 group, more than doubling relative to pre-pandemic rates (IRR = 2.11, p < 0.001). A similar approximate 2-fold increase relative to the pre-pandemic norm also occurred for the 15 to <45 and 45 to <65 groups. The greatest shift in the BC+ IR during the surge occurred for the <15 group, which increased 4-fold (IRR = 4.05, p < 0.001). Despite this alarming increase, it was less than the nearly 5-fold increase in the median iGAS IR for the <15 group during the surge. Analysis of the proportionality of the increased severe disease burden during the surge relative to the age groups revealed that only the ≥65 group was significantly disproportionately increased compared to the overall population (IRR= 1.80, p < 0.001) (Supplementary Table S3).

### Impact of the COVID-19 pandemic on absolute iGAS infection cases

As an additional measure of the potential emergence of more virulent clones with an increased capacity to cause iGAS infections in correlation with the post-pandemic surge, we next asked whether the COVID-19 pandemic resulted in a cumulative increase in infections above what would have been anticipated had the pandemic not occurred. This query was applied to both the total iGAS cases and the BC+ cases (Fig. 3A and B, respectively). Despite the marked post-pandemic surge, the cumulative number of cases observed throughout the combined pandemic and post-pandemic intervals did not differ from that expected for either total or BC+ infections if the pre-pandemic IRs had just persisted independent of the pandemic (p = 0.521 and p = 0.093, respectively). For both infection categories, the case decrease during the implementation of the COVID-19 NPIs was nearly equally offset by the transient surge in cases that occurred when the NPIs were eased. However, while the absolute number of infections did not increase, the pandemic did alter the age group distribution of the disease burden. Notably, the iGAS infection burden disproportionately impacted the <15 group (p < 0.001), increasing cases above expected, and reduced the burden on the 15 to <45 group (P < 0.001) below expected. For all but the 45 to <65 group, there were significantly fewer BC+ iGAS cases than expected based on the pre-pandemic IR norms.

**Fig. 3.**
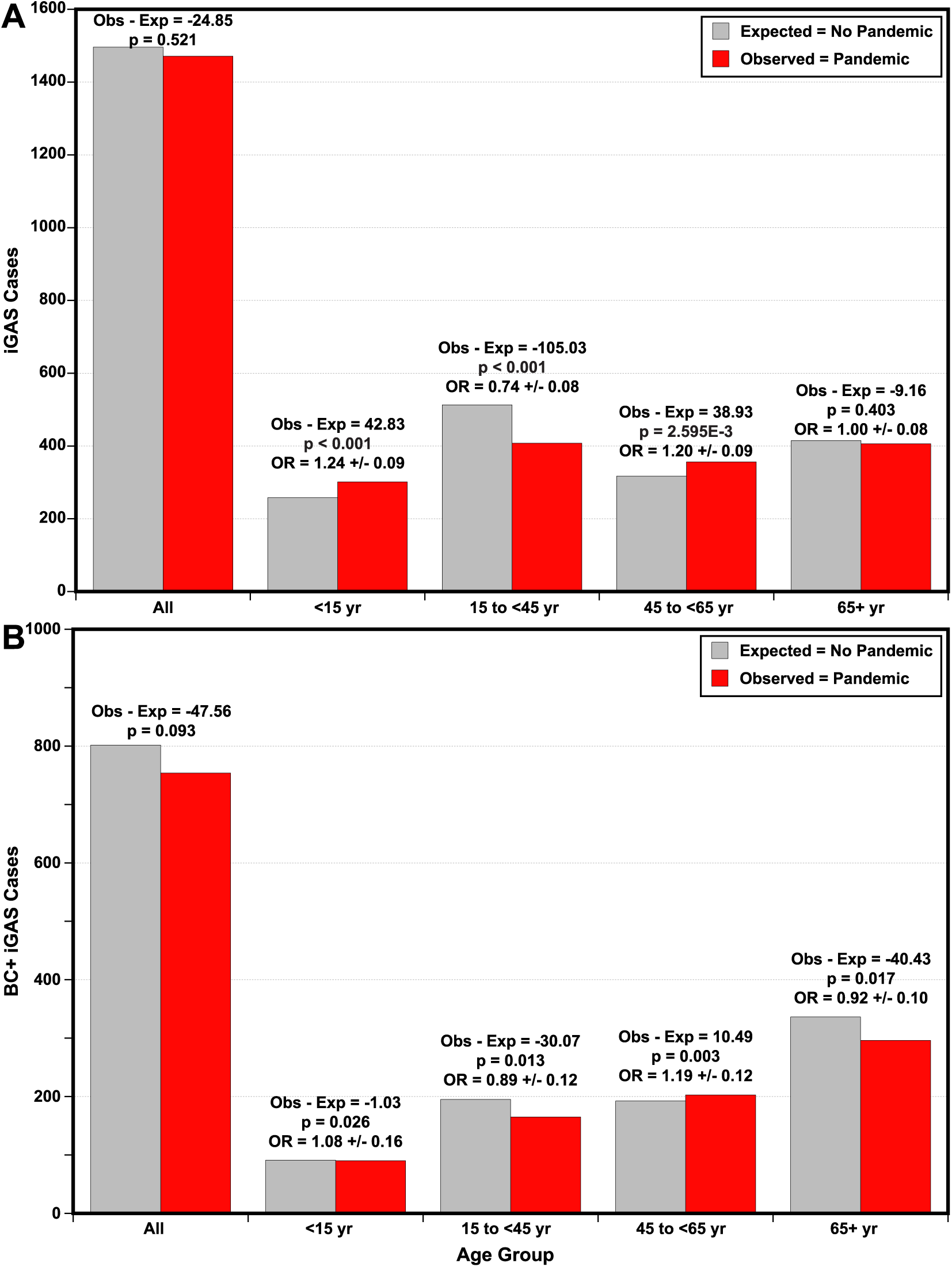
Comparison of observed and predicted invasive group A streptococcus cases for the COVID-19 pandemic and post-pandemic intervals, from April 2020 to December 2024. Expected cases across age groups were predicted using the pre-pandemic norm mean IRs and adjusted for population size. Odds ratios and p-values were determined using Chi-square goodness of fit test. (**A**) Total invasive group A streptococcus cases. (**B**) Blood culture–positive group A streptococcus cases.

### Impact of the COVID-19 pandemic on iGAS coinfection with respiratory viruses

To evaluate whether fluctuations in community respiratory virus circulation coincided with the post-pandemic surge in iGAS infections, population-level incidence rates for hospitalized influenza and RSV were compared alongside iGAS incidence over the study period (Fig. 4). Consistent with the implementation of COVID-19 NPIs, both influenza and RSV exhibited reduced activity during the pandemic interval, followed by incidence spikes overlapping with the post-pandemic iGAS surge (p = 0.045 and p = 0.012, respectively). Notably, the annual seasonal peaks for both influenza and RSV coincided directly with the late 2022 surge in iGAS incidence for both the <15 and ≥65 groups. During this period, influenza incidence rebounded across both extremes of the age spectrum, with the <15 group experiencing the highest incidence observed across the entire 11-year study period. The temporal correlation between the increase in viral respiratory infections with the post-pandemic surge in iGAS infections supports the hypothesis that viral respiratory co-infections contributed to the iGAS surge.

**Fig. 4.**
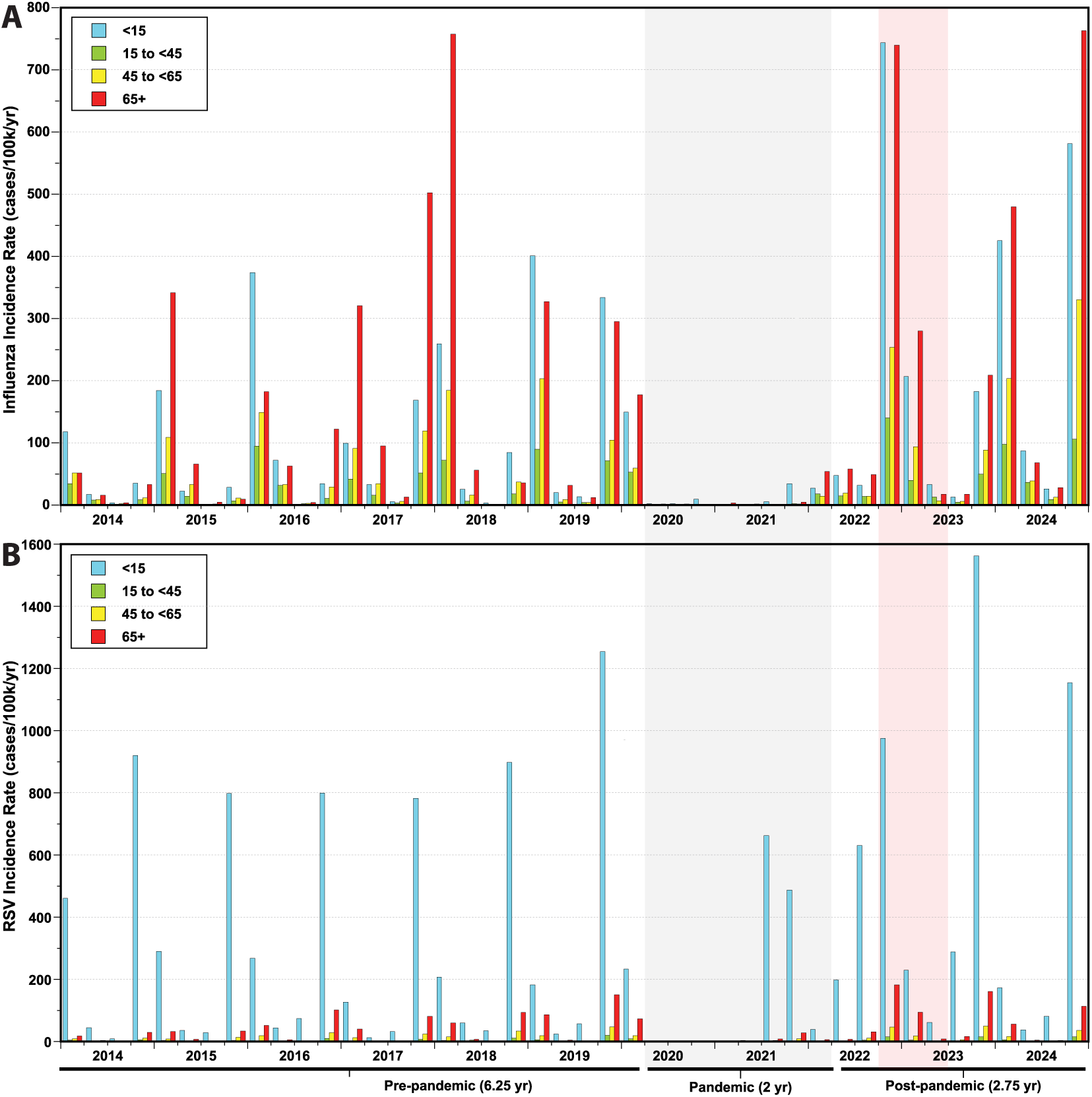
Respiratory virus quarterly incidence rates in Scotland from 2014 to 2024, stratified by age group. Pandemic and post-pandemic surge intervals are highlighted in gray and red respectively. (**A**) Influenza virus. (**B**) Respiratory syncytial virus (RSV).

### Impact of the COVID-19 pandemic on the distribution of iGAS emm types

Change in iGAS *emm* type distribution was evaluated over the study period relative to the COVID-19 pandemic (Fig. 5A). Among the 3,408 iGAS cases, 70 *emm* types were identified, with *emm* types 1, 89, 12, 3 and 76 most prevalent, accounting for 55% of the isolates. This overall distribution was stable; although the proportions varied modestly between the pre- and post-pandemic intervals, these five *emm* types remained most prevalent, accounting for 56% and 62% of the pre- and post-pandemic isolates, respectively (Fig. 5A inset). The greatest change in the *emm* type distribution occurred during the surge. While *emm*1 accounted for 23% of iGAS cases pre-pandemic, it became predominant during the surge accounting for 56% or the majority of all iGAS cases.

**Fig. 5.**
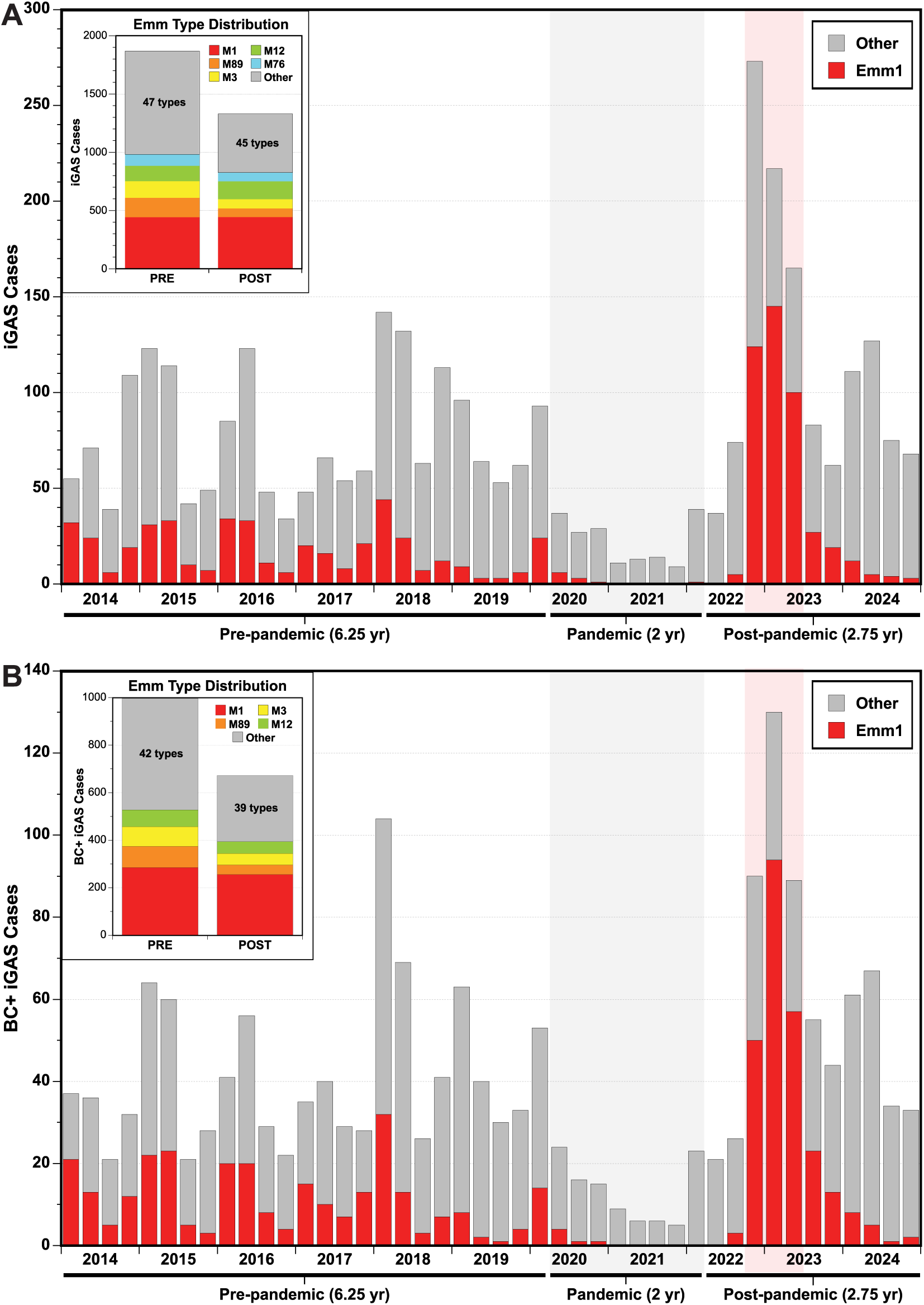
(**A**) Quarterly invasive group A streptococcus cases illustrating proportion *emm*1. The inset graph shows the abundances of the five most prevalent *emm* types (1, 12, 89, 3, and 76) during the pre-pandemic and post-pandemic intervals. (**B**) Quarterly blood culture-positive invasive group A streptococcus cases illustrating the proportion *emm*1. The inset graph shows the abundances of the four most prevalent *emm* types (1, 89, 3 and 12) during the pre-pandemic and post-pandemic intervals.

Across the 58 *emm* types identified among the 1,791 BC+ cases, the four most prevalent types 1, 89, 3, and 12, were the same as those observed among all iGAS cases (Fig. 5B). The *emm* types most prevalent for BC+ pre-pandemic remained most prevalent post-pandemic (Fig. 5B inset). While *emm*1 accounted for the majority of total iGAS infections during the surge, it was even more abundant among the BC+ cases, accounting for 65% of all BC+ isolates. That is *emm*1 caused more BC+ iGAS infections during the surge than all other *emm* types combined. Consistent with the proportion of BC+ iGAS cases being lower during the surge than pre-pandemic, the proportion of BC+ *emm*1 cases was also lower during the surge (54% ± 13%) than pre-pandemic (64% ± 13%) (p = 0.004) (Supplementary Figure S3). The reduction in the proportion of iGAS cases BC+ post-pandemic and during the surge argues against the emergence of more fit and/or virulent GAS clones in correlation with and contributing to the COVID-19 post pandemic iGAS infection surge.

### Population genomic epidemiology of the emm1 isolates

To evaluate for potential genetic changes contributing to the iGAS surge, we focused our analysis on *emm*1 isolates given their prevalence over the study period and predominance during the surge in Scotland. To permit comprehensive, genome-wide evaluation for genetic changes, both short- and long-read whole-genome sequencing was used to hybrid assemble complete closed genome sequences for 399 of 404 *emm*1 isolates studied (Supplementary Table S1).

The 404 WGS *emm*1 isolates included 232 of the 898 (25.8%) iGAS isolates, of which 122 (52.6%) were BC+; and 172 tonsillitis isolates of which 157 (91.3%) were collected during the surge. Core genome SNP phylogenetic analysis identified 16 (4.0%) isolates as M1_GLB_, 8 (2.0%) as M1_INT_, and 380 (94.0%) as M1_UK_ (Fig. 6 and Supplementary Table 4). The M1_GLB_ and M1_UK_ lineages formed separate genetically distinct clusters. In contrast, the invasive but not BC+, BC+ and tonsillitis isolates were not genetically distinct but were interspersed with each other throughout the core genome phylogeny (Fig. 6A). Similarly, although temporal structuring was evident, isolates from different COVID-19 pandemic intervals were not genetically distinct but were interspersed throughout the core genome phylogeny (Fig. 6B).

**Fig. 6.**
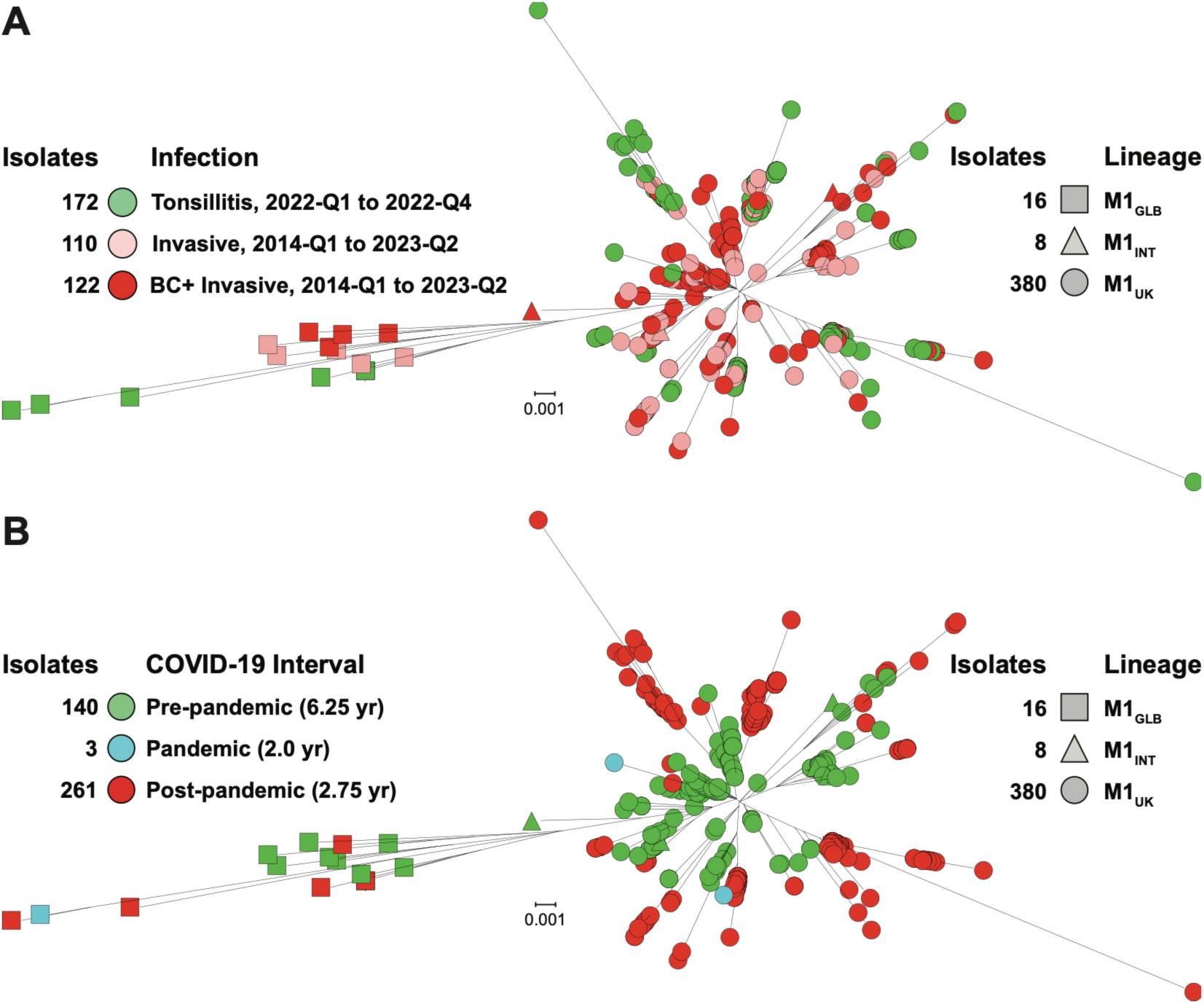
Phylogenetic relationships among 404 *emm*1 clinical isolates. Phylogeny was inferred by maximum-likelihood based on 1,710 core chromosomal single nucleotide polymorphisms (SNPs) and the trees are illustrated as radial phylograms. M1 genetic lineages (M1_GLB_, M1_INT_, and M1_UK_) are indicated by shape per the index on the right. (**A**) Isolates are color-coded by clinical infection category (tonsillitis, invasive, or blood culture-positive invasive) per the index on the left. (**B**) Isolates are color-coded by the COVID-19 pandemic interval of isolation (pre-pandemic, pandemic, or post-pandemic) per the index on the left.

### Population genomic assessment of emm1 genetic changes

We evaluated the *emm*1 genomes for polymorphisms significantly differently distributed between the pre- and post-pandemic isolates with the objective of identifying any genetic changes correlating with and potentially contributing to the post-pandemic surge in iGAS infections. The emergence of the pandemic M1_GLB_ lineage in the early 1980s was epidemiologically associated with an increase in *emm*1 iGAS infection frequency and severity (Colman et al. 1993; Musser et al. 1991; Musser JM 1998; Stevens et al. 1989). A horizontal genetic transfer (HGT)/recombination event that resulted in increased expression of the secreted virulence factors (VFs) NADase (*nga*) and streptolysin O (*slo*) was seminal to this pandemic shift (Nasser et al. 2014; Zhu et al. 2015; Stevens et al. 2000; Sumby et al. 2006). Given its potential for altering GAS pathogenesis, HGT/recombination among the 404 *emm*1 genomes was assessed using Gubbins. Recombination was found minimal among the 404 *emm*1 genomes (Supplementary Fig. S4). Recombination blocks (RBs) were inferred at 39 loci genome-wide for the cohort of which only three involved more than eight isolates. These three loci were for the pilus regulator *rofA* (MGAS2221_0149, 3 SNPs in 388 isolates), an IS1548 family transposase (MGAS2221_1549, 3-6 SNPs in 391 isolates), and the streptococcal inhibitor of complement secreted VF *sic* (MGAS2221_1699, 8-39 SNPs in 75 isolates). The three SNPs in *rofA* are part of the 27 SNPs defining the M1_UK_ lineage and were present in all M1_INT_ and M1_UK_ isolates, but none of the M1_GLB_ lineage isolates. None of the 39 RBs were significantly non-randomly distributed with respect to the pre- and post-pandemic intervals. Thus, no evidence was found supporting HGT/recombination as correlating with and contributing to the post-pandemic iGAS surge.

We next conducted a genome-wide association study (GWAS) of the 380 M1_UK_ lineage isolates relative to the pandemic intervals using PYSEER. The GWAS was done at two levels: 1) with unitigs to evaluate for small genetic polymorphisms (primarily SNPs and indels) genome-wide; and 2) with gene content to evaluate for larger differences in the accessory portion of the pangenome (composed primarily of mobile genetic elements, such as phages and integrative-conjugative elements) variably present among the individual isolates. For any genetic polymorphism to be meaningfully contributing to the surge it would need to be strongly differentially distributed between the 124 pre-pandemic and 254 post-pandemic M1_UK_ genomes -- that is it would need to have an intermediate allele frequency (AF) being neither largely present nor largely absent among all the genomes.

Among 14,573 unitigs (collapsed overlapping 31-mer sequences encompassing one or more polymorphisms such as SNPs) evaluated, the AF was strongly bimodal with the unitigs being either mostly present (AF ≥ 0.9, *n* = 8,852) or mostly absent (AF ≤ 0.1, *n* = 5,590) in all 380 genomes (Fig.7). Only 92 unitigs were significantly non-randomly distributed pre- and post-pandemic, of which only four had an intermediate AF (> 0.3 and < 0.7). None of these four unitigs were absent among >50% of the pre-pandemic isolates and present among >50% of the post-pandemic isolates, or vice-versa. Moreover, none of the four involved VFs or VF regulators. Similarly, among 2,010 genes in the M1_UK_ cohort pangenome, only17 were significantly non-randomly distributed, and only one had an intermediate AF (Fig.7). This gene encoding a small hypothetical protein (∼50aa), was detected in 61 pre-pandemic and 38 post-pandemic genomes. This gene is commonly not unique in GAS genomes being found 3’ adjacent to IS1548 transposases and is frequently disrupted and unannotated. These pangenome gene content results show that the post-pandemic M1_UK_ isolates as a group have not acquired a new MGE (phage or ICE) encoding additional VFs. This finding is consistent with an analysis of the closed genomes which found that the vast majority, 346 (85.6%) of the 404 *emm*1 genomes, have the same complement of three phages encoding VFs as are present in the genome of the M1_GLB_ reference strain MGAS2221 (phages 2221.1 encoding SpeA2, 2221.2 encoding Spd3, and 2221.3 encoding SdaD). Consistent with the finding that no variably present genes had a distribution indicating the broad acquisition of a new MGE temporally correlating with the surge, analysis of antimicrobial resistance gene content found that only 2 of the 404 *emm*1 genomes possessed known resistance determinants—specifically the macrolide resistance gene *erm(B)* and the tetracycline resistance gene *tet(M)*. Therefore, the GWAS analyses found no evidence for acquisition of novel polymorphisms or virulence factors, nor a shift in antibiotic resistance correlating with and contributing to the post-pandemic iGAS surge.

**Fig. 7.**
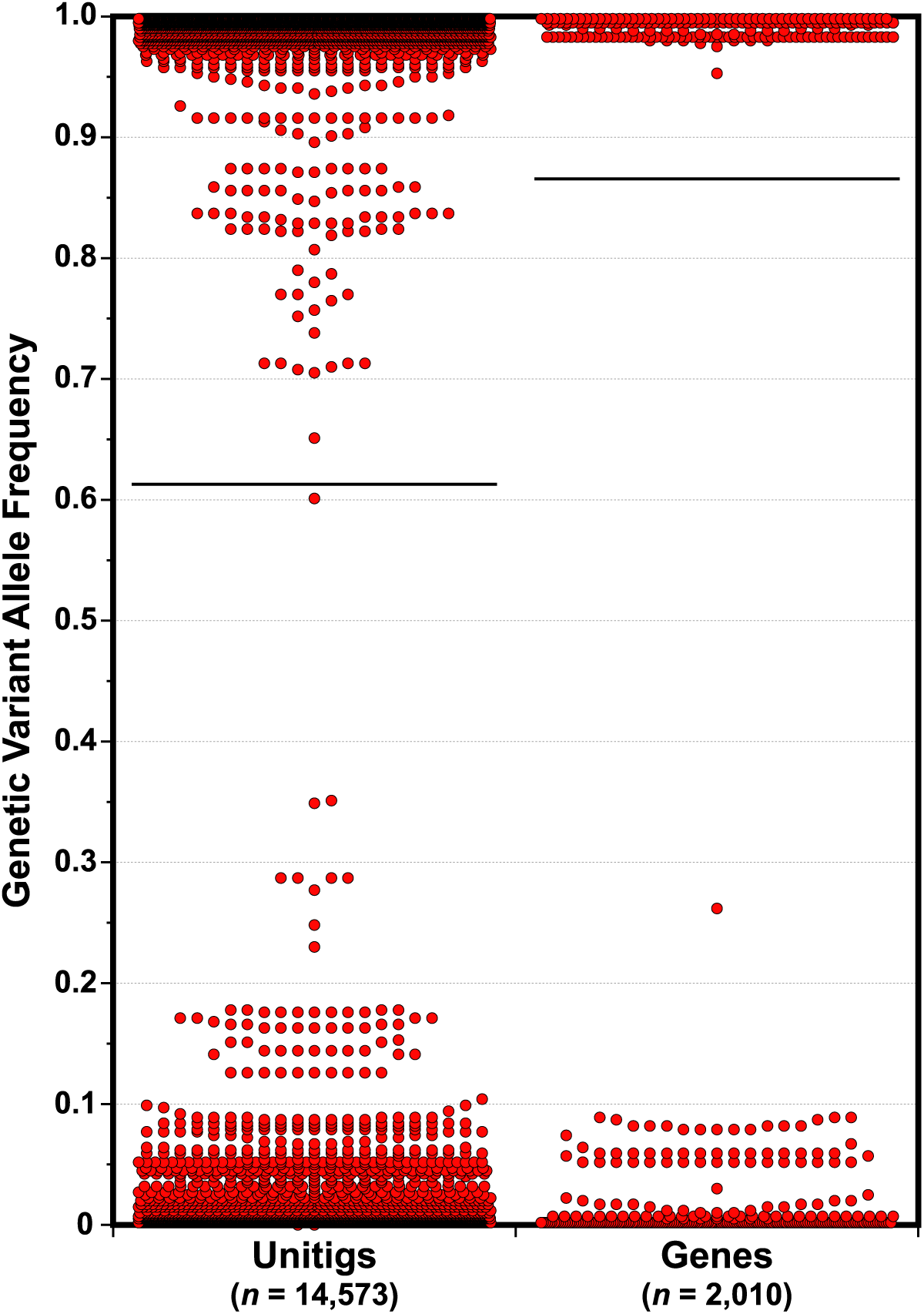
PYSEER genome-wide association study (GWAS) allele frequencies. Allele frequencies (AF) of genetic variants identified among the 380 M1_UK_ clinical isolates are plotted. The AF distribution for 14,573 unitigs, representing small genetic polymorphisms (SNPs and indels) evaluated genome-wide, is shown on the left, and the distribution for 2,010 pangenome gene clusters is on the right. The distribution of both unitigs and genes is strongly bimodal (most variants having an AF ≤ 0.1 or ≥ 0.9), demonstrating a lack of intermediate-frequency genetic variants that would be expected if a novel *emm*1 clone had emerged correlating with and contributing to the post-pandemic surge.

Some investigations have noted expansion of *emm*3.93 GAS isolates having variant chromosomal architectures (inversions and rearrangements) in correlation with or closely following the post-pandemic iGAS surge and speculated that these structural variants (SVs) may represent the emergence of more fit and/or virulent clones (Davies et al. 2025; Pilar et al. 2026). Such large SVs are particularly relevant to the *emm*1 type as M1_GLB_ lineage reference strains MGAS5005 and MGAS2221 differ in chromosomal architecture consistent with two inversion events symmetric about the origin of replication. Notably one of these inversion events changes gene content downstream adjacent to global virulence regulator. Mga. Alignment analysis of the 399 closed *emm*1 genomes for large SVs revealed that 395 of the 399 closed genomes were collinear with MGAS2221 (Supplementary Table S4). Because the hybrid genome assemblies represent a consensus of the underlying sequence reads, this indicates that the MGAS2221 conformation represents the predominant architecture within these cultures. However, it is important to understand that the consensus assembly process does not address the frequency at which such chromosomal rearrangement events occur, and therefore any given isolate culture may actually contain multiple SV chromosomal architectures. Despite these caveats, the hybrid assemblies provide no evidence for a shift in *emm*1 chromosomal architecture correlating with or contributing to the post-pandemic iGAS infection surge.

## Discussion

While post-pandemic iGAS surges in several countries were reported to predominantly affect children (Abo et al. 2023; Guy et al. 2023; Nygaard et al. 2024), our findings indicate that in Scotland, as reported in Germany (Goldberg-Bockhorn et al. 2024; Singer et al. 2024), both children and older adults were disproportionately impacted, consistent with these age groups representing the most immunologically vulnerable populations (McGregor et al. 2023). The observation that incidence initially peaked in children in late 2022, followed closely by a later peak in older adults in early 2023, suggests age-dependent transmission dynamics, in which increased transmission in pediatric populations preceded and contributed to subsequent exposure in older adults. Reopening of schools following the easing of the COVID-19 pandemic NPIs in the summer 2022 likely increased the exposure levels of children not only to GAS but also to respiratory viruses known to predispose for iGAS disease (Herrera, Huber, and Chaussee 2016), which also resurged during the post-pandemic period in Scotland and other settings (Lassoued et al. 2023).

Although blood culture positive iGAS IRs increased for all age groups during the post-pandemic surge, the proportion of iGAS cases BC+ (i.e. the iGAS BC+ rate) was lower than during the pre-pandemic period. This finding argues against the hypothesis that the surge was characterized by a shift towards greater disease severity and was contributed to by the emergence of more virulent GAS clones. Several factors may have contributed to the reduced proportion of bacteremia cases during the surge, including increased reporting of less severe iGAS presentations during a period of heightened awareness and the disproportionate increase in pediatric cases, an age group significantly less likely than older adults to develop bacteremia. Although such factors may have introduced bias into our estimates of bacteremia risk, our findings remain inconsistent with a substantial increase in disease severity during the surge. This interpretation is supported by another surveillance study in Scotland, which reported an overall increase in severe iGAS infections in children during 2022–2023 but found mortality rates comparable to those observed before the pandemic (Holdstock et al. 2025). Similar findings have been reported in the Netherlands and Denmark, where increases in iGAS incidence during the post-pandemic period were not accompanied by significant changes in case-fatality rates (Davies et al. 2025; Johannesen et al. 2023; van Kempen et al. 2025). In Scotland throughout the study surveillance period individuals aged ≥65 years experienced the highest burden of bacteremia disease, whereas children aged <15 years, despite showing the largest increase in overall iGAS incidence, were not disproportionately affected by bacteremia. These age-specific differences are more consistent with variation in immunity related host susceptibility and exposure patterns than with a uniform increase in pathogen virulence across demographic groups.

The overall number of iGAS and BC+ cases across the pandemic and post-pandemic periods did not differ significantly from that expected based on pre-pandemic incidence. That is the COVID-19 pandemic did not result in an overall increase in iGAS cases above and beyond what would have been anticipated based on the pre-pandemic IRs. This finding suggests that the marked reduction in iGAS cases during the pandemic was equally offset by a transient compensatory increase during the post-pandemic surge. These findings are broadly consistent with the concept of “immunity debt”. The implementation of NPIs to reduce SARS-CoV-2 transmission reduced exposure to a range of respiratory pathogens, decreasing naturally acquired immunity at the population level (Burrell, Saravanos, and Britton 2025; Cohen et al. 2021; Dokal et al. 2025; Flamant et al. 2025; Lorenz et al. 2025). The subsequent lifting of these measures in 2022 likely resulted in increased exposure to multiple infectious agents in a population with reduced immunity causing a transient infection surge until population immunity was re-established through the restoration of pre-pandemic exposure levels. A similar pattern was observed for influenza and RSV, both risk factors for GAS disease (Burrell, Saravanos, and Britton 2025; Dokal et al. 2025; Flamant et al. 2025; Lorenz et al. 2025). Both viral infections showed a marked decrease during the pandemic, followed by peaks coinciding with the iGAS surge in late 2022, consistent with observations reported in other countries (Burrell, Saravanos, and Britton 2025; Flamant et al. 2025). While these temporal associations do not establish causality, they support the hypothesis that increased circulation of respiratory viruses likely contributed to the post-pandemic resurgence of iGAS disease.

The COVID-19 pandemic did not result in a shift in the distribution of the most prevalent *emm* types in Scotland. The *emm* types that were the most prevalent causes of iGAS infection prior to the pandemic were the same types most prevalent following the pandemic. Although the M1_UK_ lineage was the predominant cause of iGAS infection during the post-pandemic surge in Scotland, it was not associated with higher rates of bacteremia than *emm*1 isolates pre-pandemic, arguing against a shift to increased virulence in correlation with the post-pandemic surge in iGAS infections. This is consistent with the finding that, despite the M1_UK_ lineage being commonly described as "hypervirulent", mortality was found to not differ significantly between the M1_UK_ and M1_GLB_ lineages among 1,356 *emm*1 iGAS cases investigated in the United Kingdom (Li et al. 2025). Phylogenetic analysis indicated that disease severity among *emm*1 isolates was not associated with distinct phylogenetic clusters, consistent with findings reported elsewhere (Vieira et al. 2024). Similarly, in Scotland the *emm*1 post-pandemic isolates were distributed across multiple branches of the phylogeny, with no evidence for the expansion of a single dominant clone. Analogous dynamics have been reported in other settings, where multiple *emm*1 sublineages appeared to have expanded during the post-pandemic surge (Morris et al. 2025; Rumke et al. 2024; Vieira et al. 2024). Indeed, genomic and epidemiological surveillance from the United States illustrates that the post-pandemic iGAS surge occurred independently of the M1_UK_ lineage expanding and becoming predominant. National US surveillance from 2019 to 2021 demonstrated that M1_UK_ accounted for only 11% of *emm*1 iGAS infections (Li et al. 2023). During the height of the 2022–2023 surge, high-resolution genomic analysis of isolates from North Carolina revealed that the predominant *emm*1 strains belonged to distinct sub-lineages rather than M1_UK_ (Huang et al. 2024). This is further supported by epidemiological data from Texas, Colorado, and Minnesota, where the surge was driven heavily by *emm*12 or a polyclonal mix of lineages (Aboulhosn et al. 2023; Barnes et al. 2023; Dabaja-Younis et al. 2025). In the aggregate, these findings demonstrate that the association between the expansion of the M1_UK_ lineage and the post-COVID-19 pandemic surge in iGAS infections found in some investigations (Beres et al. 2024; Gouveia et al. 2023; Guy et al. 2023; Rodriguez-Ruiz et al. 2023; Rumke et al. 2024; Vieira et al. 2024) is not common to all studies, arguing against the simple explanation that the post-pandemic surge in iGAS infections was driven by the emergence and expansion of a more virulent clone.

In an effort to explain the unprecedented post-pandemic surge in iGAS infections, multiple recent investigations have speculated on the emergence and expansion of a more fit, transmissible, or hypervirulent clone, such as the M1_UK_ or M1_DK_ lineages (de Gier et al. 2024; Johannesen et al. 2023; Vieira et al. 2024). However, in virtually all these reports, the comprehensive population-level genomic analyses required to rigorously test this emergent more virulent clone hypothesis have been lacking. Previous studies have predominantly relied on short-read sequencing platforms or allele-specific PCR (Zhi et al. 2023), methodologies that produce incomplete genomic characterizations that cannot definitively resolve both small (SNPs and indels) nor large (gene and MGE content nor chromosomal SVs) genetic variants across the population. Furthermore, these speculations regarding increased pathogen fitness have not been corroborated by animal models of infection demonstrating an experimental phenotypic increase in virulence for the surge isolates (Vieira et al. 2024). We have addressed this gap in complete closed genome data for the evaluation of the emergence of genetic variants in association with the post-pandemic iGAS surge with the largest set of hybrid WGS collection of clinical isolates of any single human bacterial pathogen conducted to date. By independently resolving the complete, closed chromosomal architecture for 399 clinical isolates using hybrid assembly, our investigation allowed a robust determination of all genetic polymorphisms, from small (i.e. SNPs and indels) to large (i.e. gene/MGE content and chromosomal SVs) comprehensively genome wide. Regardless of the size, no polymorphisms were found consistent with the emergence of a novel *emm*1 clone in association with the post-pandemic surge in *emm*1 iGAS infections in Scotland. When considered alongside the epidemiological findings, these genomic data support the interpretation that the post-pandemic surge reflected expansion of a pre-existing *emm*1 population under conditions of increased host susceptibility, rather than the emergence of a more fit and/or virulent *emm*1 clone.

The genomic component of this study by design was skewed in focus on the pediatric cohort which was disproportionately impacted by the iGAS post-pandemic surge. Although other age groups are less well represented, the large number of *emm*1 isolates analyzed over the study period is likely representative of GAS infections irrespective of patient age. Additional limitations of this study include the lack of linkage to detailed clinical and socioeconomic data, which precluded evaluation of changes in clinical presentation and established risk factors such as socioeconomic status and access to health care, and may limit interpretation of transmission dynamics across population subgroups. Finally, the true burden of iGAS disease may be underestimated due to incomplete case ascertainment and isolate submission.

In conclusion, this study provides strong evidence that in Scotland the post-pandemic surge in iGAS cases was not driven by the emergence of more virulent GAS clone/s. Instead, our data support the resurgence of pre-existing strains in the context of increased population susceptibility following the pandemic. The disproportionate burden observed in children and older adults highlights the role of age-specific susceptibility and transmission dynamics. These findings help to clarify those factors that contributed to the post-pandemic surge in iGAS infections and provide an evidence base to inform targeted public health strategies and prioritize interventions aimed at protecting high-risk populations.

## Data Availability

All data produced in the present study are available upon reasonable request to the authors, and all whole genome sequencing data have been submitted to the NCBI SRA under Bioproject accessions PRJNA1076228 and PRJNA1459430.

## Declaration of generative AI in the manuscript preparation

During the preparation of this work the artificial intelligence (AI) tool NotebookLM was used to facilitate drafting, revising, and structuring the epidemiological and genomic data narratives, and Gemini to generate R code for statistical tests, including the Poisson and logistic regression analyses. After using these tools, the authors reviewed and edited all AI generated content and take full responsibility for the content of the published article.

## Data availability

Genomic data for the 404 *emm*1 isolates generated for this investigation were deposited into the National Center for Biologic Information Sequence Read Archive under Bioproject accessions PRJNA1076228 and PRJNA1459430.

## Acknowledgements

We acknowledge the Scottish diagnostic laboratories in referring iGAS isolates to the Reference Laboratory. All staff from the Scottish Reference Laboratory, Glasgow for processing isolates. To Morris Muzyumba and Eisin McDonald Public Health Scotland for discussions and data concerning iGAS case epidemiology. Additionally, we acknowledge the members of the Houston Methodist genomics team: Alma Amaya, Regan Mangham, Mattew Ojeda Saavedra, Jordan Pachua, and Sindy Pena.

