## Supplemental Tables S2 and S3 for "Clinical and population genomic epidemiology of invasive group A streptococcus in Scotland, 2014-2024"

**Supplementary Table S2.**

Proportionality of invasive Group A Streptococcus (iGAS) relative risk by age group during the pre-pandemic and post-pandemic surge intervals.

| COVID-19 Interval | | All ages | <15 yr | 15 to <45 yr | 45 to <65 yr | ≥65 yr |
|---|---|---|---|---|---|---|
| Pre-pandemic 2014-Q1 to 2020-Q1 | IR [IQR] | 4.7 [3.9-8.2] | 5.6 [3.8-8.0] | 4.2 [3.2-6.7] | 4.2 [2.7-5.6] | 7.9 [5.1-9.6] |
| | IRR (95% CI) | -- | 1.13 (1.01-1.26) | 0.92 (0.85-1.01) | 0.77 (0.70-0.86) | 1.38 (1.25-1.52) |
| | p-value | -- | 0.039 | 0.073 | <0.001 | <0.001 |
| Surge 2022-Q4 to 2023-Q2 | IR [IQR] | 15.8 [13.9-17.9] | 24.5 [20.4-39.6] | 9.1 [8.6-9.8] | 12.6 [12.3-12.9] | 21.5 [18.8-23.3] |
| | IRR (95% CI) | -- | 1.97 (1.70-2.29) | 0.60 (0.51-0.71) | 0.79 (0.66-0.93) | 1.29 (1.10-1.51) |
| | p-value | -- | <0.001 | <0.001 | 0.006 | 0.002 |
| | IRR (Surge/Pre-pandemic) | 3.36 | 4.38 | 1.63 | 3.00 | 2.72 |

The median incidence rate (IR) of iGAS for individual age cohorts (<15, 15 to <45, 45 to <65, and ≥65 years) was compared against the median IR across all age groups combined. Incidence rate ratios (IRR) with 95% confidence intervals (CI) and statistical significance (p-values) were calculated using a generalized linear model for Poisson regression. The IRRs represent the relative risk of iGAS infection for a specific age group compared to the overall population median during both the pre-pandemic historic norm and the post-pandemic surge intervals.

**Supplementary Table S3.**

Proportionality of blood culture positive (BC+) Invasive Group A Streptococcus (iGAS) relative risk by age group during the pre-pandemic and post-pandemic surge intervals.

| COVID-19 Interval | | All ages | <15 yr | 15 to <45 yr | 45 to <65 yr | ≥65 yr |
|---|---|---|---|---|---|---|
| Pre-pandemic 2014-Q1 to 2020-Q1 | IR [IQR] | 2.7 [2.2-3.9] | 1.9 [1.4-3.3] | 4.2 [3.2-6.7] | 4.2 [2.7-5.6] | 7.9 [5.1-9.6] |
| | IRR (95% CI) | -- | 0.74 (0.62-0.90) | 0.92 (0.85-1.01) | 0.77 (0.70-0.86) | 1.38 (1.25-1.52) |
| | p-value | -- | 0.002 | 0.073 | <0.001 | <0.001 |
| Surge 2022-Q4 to 2023-Q2 | IR [IQR] | 6.6 [6.5-8.0] | 8.2 [8.2-9.8] | 3.9 [3.7-4.0] | 6.5 [5.8-8.1] | 12.0 [11.0-15.3] |
| | IRR (95% CI) | -- | 1.23 (0.93-1.63) | 0.51 (0.39-0.68) | 0.94 (0.73-1.20) | 1.80 (1.45-2.24) |
| | p-value | -- | <0.142 | <0.001 | 0.627 | <0.001 |
| | IRR (Surge/Pre-pandemic) | 2.44 | 4.32 | 2.05 | 2.82 | 2.03 |

The median incidence rate (IR) of severe BC+ iGAS infections for individual age cohorts (<15, 15 to <45, 45 to <65, and ≥65 years) was compared against the median IR across all age groups combined. Incidence rate ratios (IRR) with 95% confidence intervals (CI) and statistical significance (p-values) were calculated using a generalized linear model for Poisson regression. The IRRs represent the relative risk of a severe BC+ iGAS infection for a specific age group compared to the overall population median during both the pre-pandemic historic norm and the post-pandemic surge intervals.
