## Supplemental Figs S1-to-S4 for "Clinical and population genomic epidemiology of invasive group A streptococcus in Scotland, 2014-2024"

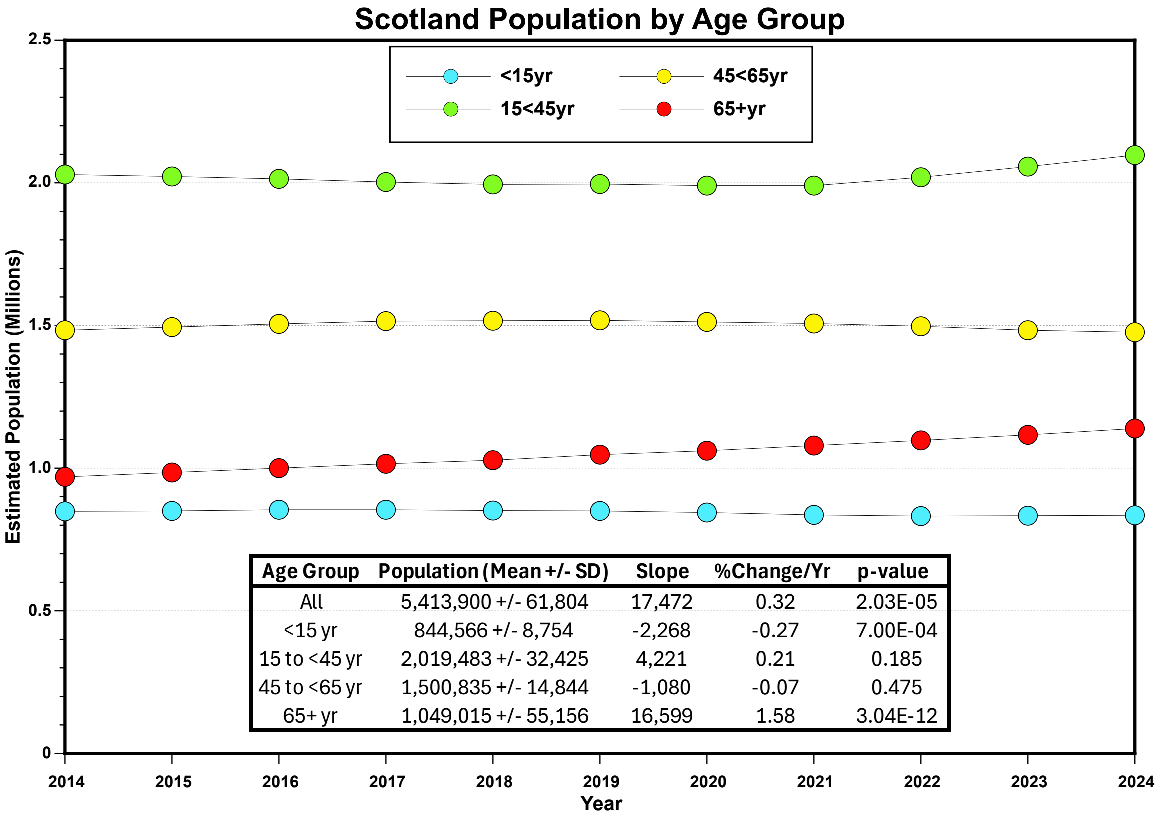


| Age Group | Population (Mean +/- SD) | Slope | %Change/Yr | p-value |
|---|---|---|---|---|
| All | 5,413,900 +/- 61,804 | 17,472 | 0.32 | 2.03E-05 |
| <15 yr | 844,566 +/- 8,754 | -2,268 | -0.27 | 7.00E-04 |
| 15 to <45 yr | 2,019,483 +/- 32,425 | 4,221 | 0.21 | 0.185 |
| 45 to <65 yr | 1,500,835 +/- 14,844 | -1,080 | -0.07 | 0.475 |
| 65+ yr | 1,049,015 +/- 55,156 | 16,599 | 1.58 | 3.04E-12 |

**Supplementary Figure S1.** Scotland population size and linear regression trend analysis, 2014–2024. The mean population size ± standard deviation for all ages and stratified by age cohorts (<15, 15 to <45, 45 to <65, and ≥65 years) is presented for the 11-year surveillance period. Linear regression trend analysis indicates the annual population slope, percentage change per year, and statistical significance (p-value). The data demonstrates that the overall Scottish population and individual age groups remained highly stable across the study period, not varying in correlation with the COVID-19 pandemic intervals. Population sizes are mid-year population estimates obtained from the National Records of Scotland.

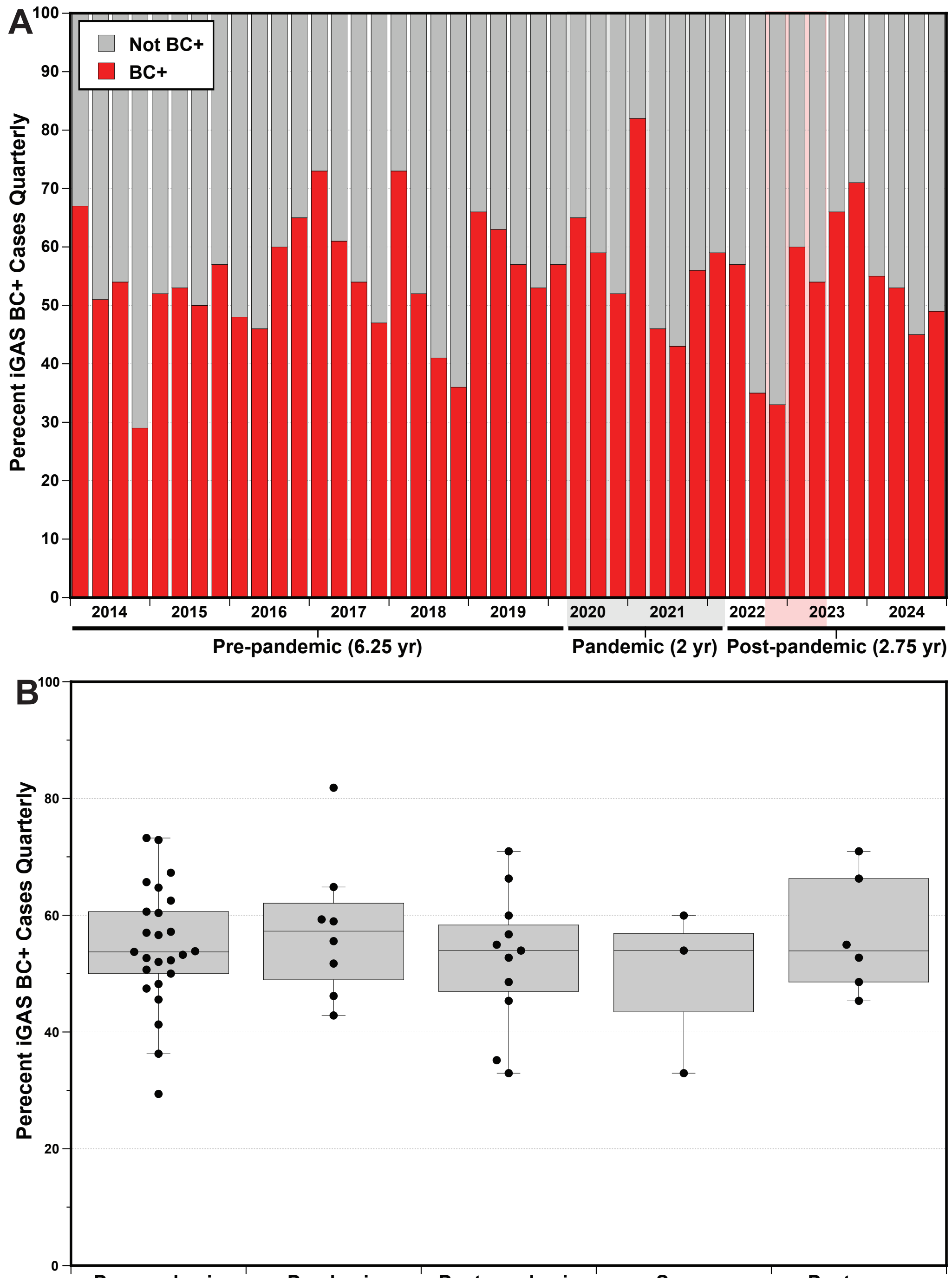


| | Pre-pandemic | Pandemic | Post-pandemic | Surge | Post-surge |
|---|---|---|---|---|---|
| **Cases:** | 1937 | 179 | 1292 | 655 | 381 |
| **Quarters:** | 25 | 8 | 11 | 3 | 6 |
| **%BC+ Median [IQR]:** | 53.7 [50.0-60.6] | 57.3 [50.3-60.7] | 53.9 [46.9-58.3] | 53.9 [43.5-56.9] | 53.9 [49.6-59.4] |
| **Odds Ratio (95% CI):** | **LR:** m = 0.181 | 1.20 (0.88-1.64) | 0.88 (0.76-1.01) | 0.77 (0.65-0.92) | 1.10 (0.90-1.33) |
| **p-value:** | **LR:** p = 0.540 | 0.247 | 0.068 | 0.005 | 0.347 |

**Supplementary Figure S2.** Proportion of invasive Group A Streptococcus cases testing blood culture-positive in Scotland, 2014–2024. (A) The quarterly percentage of iGAS cases that were blood culture-positive (BC+). (B) The quarterly percentage of iGAS cases that were BC+ stratified by COVID-19 pandemic-related intervals. Boxes display the median and interquartile range (IQR). Slope (m), Odds ratios (OR) with 95% confidence intervals (CI) relative to the pre-pandemic normal, along with the p-values determined by linear and logistic regression, are presented. The odds of an iGAS case being BC+ were significantly lower only during the post-pandemic surge interval where it was decreased by 23% relative to pre-pandemic.

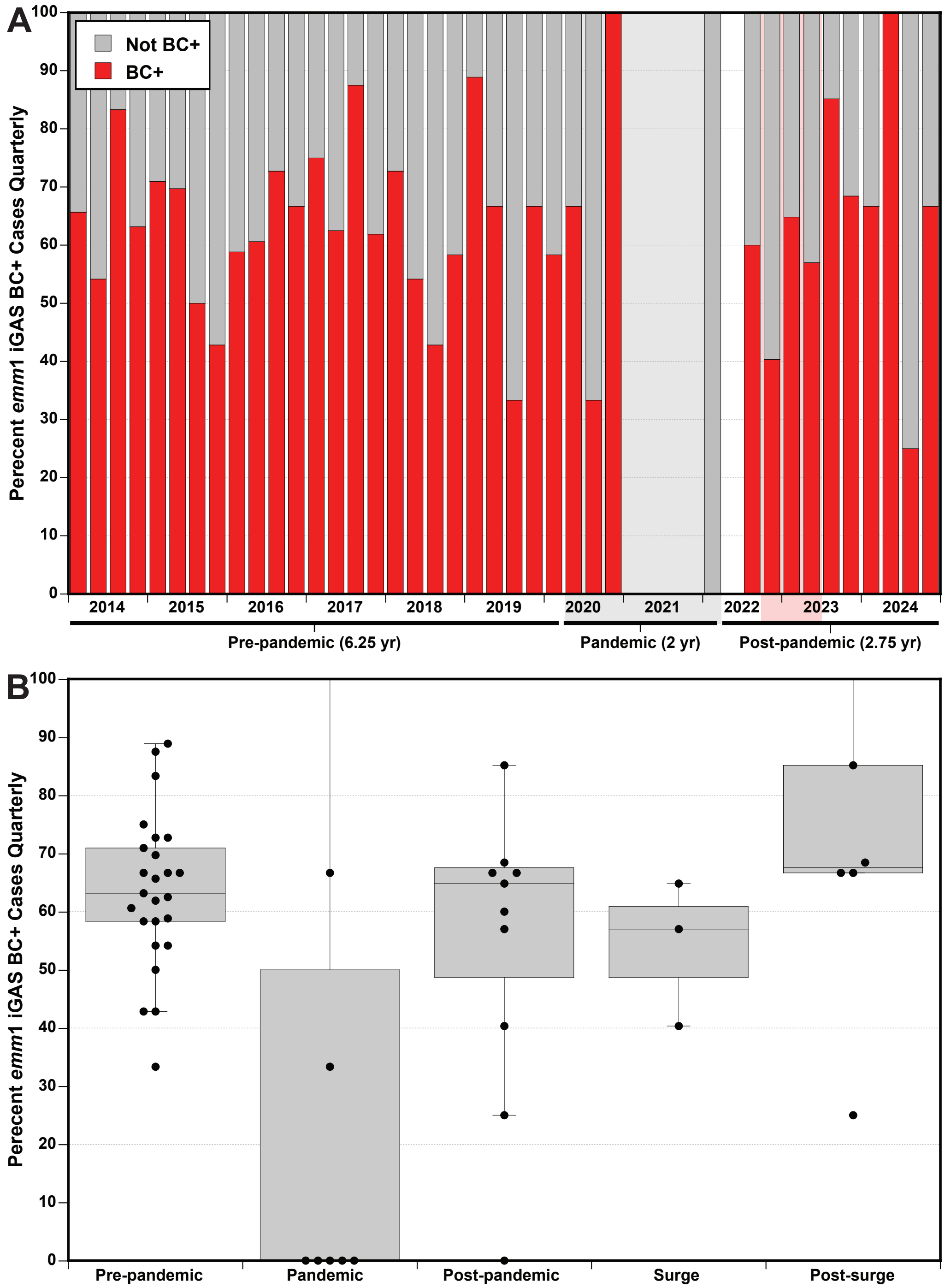


| | Pre-pandemic | Pandemic | Post-pandemic | Surge | Post-surge |
|---|---|---|---|---|---|
| **Cases:** | 443 | 11 | 444 | 369 | 24 |
| **Quarters:** | 25 | 8 | 11 | 3 | 6 |
| **%BC+ Median [IQR]:** | 63.2 [58.3-71.0] | 0.0 [0.0-41.7] | 64.8 [48.7-67.5] | 57.0 [48.7-60.9] | 67.5 [66.3-66.7] |
| **Odds Ratio (95% CI):** | **LR:** m = 0.197 | 0.67 (0.20-2.34) | 0.75 (0.58-0.99) | 0.66 (0.50-0.88) | 1.60 (0.92-1.33) |
| **p-value:** | **LR:** p = 0.604 | 0.507 | 0.042 | 0.004 | 2.90 |

**Supplementary Figure S3.** Proportion of invasive Group A Streptococcus *emm*1 cases testing blood culture-positive in Scotland, 2014–2024. (A) The quarterly percentage of iGAS *emm*1 cases that were blood culture-positive (BC+). (B) The quarterly percentage of iGAS *emm*1 cases that were BC+ stratified by COVID-19 pandemic-related intervals. Boxes display the median and interquartile range (IQR). Slope (m), Odds ratios (OR) with 95% confidence intervals (CI) relative to the pre-pandemic normal, along with the p-values determined by linear and logistic regression, are presented. The odds of an *emm*1 iGAS case being BC+ were significantly lower during the post-pandemic surge interval, where it was decreased by 34% relative to pre-pandemic.

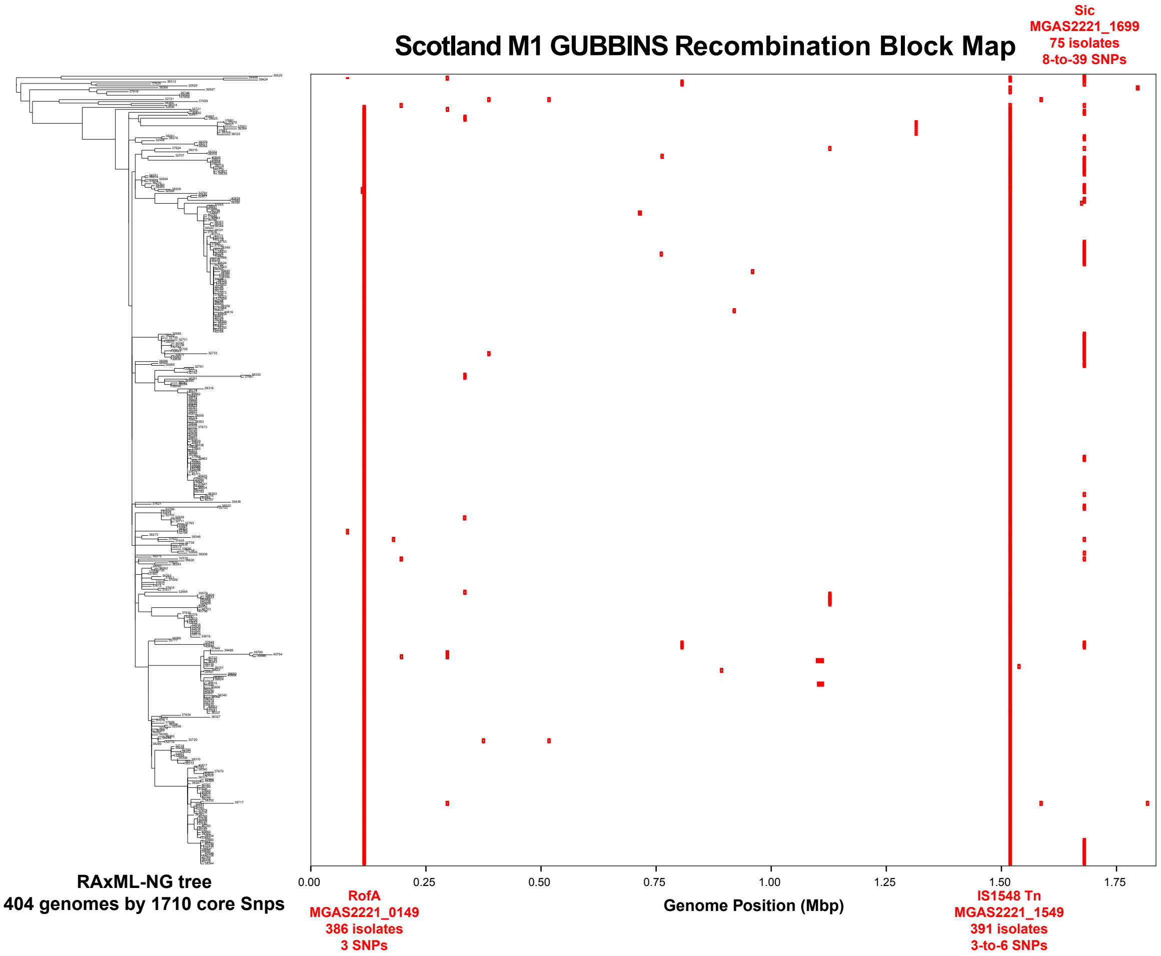


**Supplementary Figure S4.** Gubbins core genome recombination block map for 404 *emm*1 clinical isolates. The left panel displays the maximum-likelihood phylogenetic tree inferred from 1,710 core chromosomal single nucleotide polymorphisms (SNPs) determined relative to the reference MGAS2221 genome. The right panel illustrates the distribution of predicted recombination blocks (RBs), depicted as red horizontal bars, mapped against the reference genome position. Overall inferred recombination was minimal; of the 39 predicted core genome RBs, only three involved more than eight isolates. These three loci correspond to the pilus regulator *rofA* (MGAS2221_0149), an IS1548 family transposase (MGAS2221_1549), and the streptococcal inhibitor of complement secreted virulence factor *sic* (MGAS2221_1699). None of the 39 RBs were significantly non-randomly distributed between the pre-pandemic and post-pandemic intervals, indicating that recombination was not a contributing factor to the post-pandemic iGAS surge.
